# HIV burden and the global fast-track targets progress among pregnant women in Tanzania calls for intensified case finding: Analysis of 2020 antenatal clinics HIV sentinel site surveillance

**DOI:** 10.1101/2023.05.07.23289635

**Authors:** Erick Mboya, Mucho Mizinduko, Belinda Balandya, Jeremiah Mushi, Amon Sabasaba, Davis Elias Amani, Doreen Kamori, George Ruhago, Prosper Faustine, Werner Maokola, Veryeh Sambu, Mukome Nyamuhagata, Boniphace S. Jullu, Amiri Juya, Joan Rugemalila, George Mgomella, Sarah Asiimwe, Andrea B. Pembe, Bruno Sunguya

## Abstract

**Background:** For successful HIV response, updated information on the burden and progress towards elimination targets are required to guide programmatic interventions. We used data from the 2020 HIV sentinel surveillance to update on the burden, HIV status awareness, ART coverage, and factors associated with HIV infection among pregnant women in Tanzania mainland.

**Methodology:** We conducted the ANC surveillance in 159 ANC sites from all 26 regions of Tanzania’s mainland from September to December 2020. This cross-sectional study included all pregnant women (≥15 years) on their first ANC visit in the current pregnancy during the survey period. Routine HIV counseling and testing were done at the facility. Multivariable logistic regression model was used to examine factors associated with HIV infections.

**Results:** A total of 38,783 pregnant women were enrolled (median age (IQR) =25 (21–30) years). HIV prevalence was 5.9% (95%CI: 5.3% - 6.6%), ranging from 1.9% in Manyara region to 16.4% in Njombe region. Older age, lower and no education, not being in marital union, and living in urban or semi-urban areas was associated with higher odds of HIV infection. HIV status awareness among women who tested positive was 70.9% (95% CI: 67.5%- 74.0%). ART coverage among those aware of their status was 91.6% (86.5%- 94.9%). Overall, 66.6% (95% CI: 62.4%- 70.6%) of all pregnant women tested positive for HIV knew their HIV status and were on ART.

**Conclusion:** HIV is increasingly prevalent among pregnant women in Tanzania especially among older, those with lower or no formal education, those outside marital union and pregnant women living in urban and semi-urban areas. Behind the global fast-target to end HIV/ AIDS, about a third of pregnant women living with HIV initiating ANC were not on ART. Interventions to increase HIV testing and linkage to care among women of reproductive age should be intensified.

## Introduction

About 19.7 million women above 15 years were living with HIV in 2021, and about 1.3 million become pregnant each year (1,2). The risk of transmission of HIV from mother to child ranges from 15% to 45% during pregnancy, labour, delivery and breastfeeding, if proper preventive measures are not taken (3,4). In 2020 alone, about 160,000 children had newly acquired HIV principally through vertical transmission, and about 1.8 million children were living with HIV worldwide (1). The African region is the most affected region with 25.6 million people living with HIV (PLHIV) and about 60% of the global new HIV infections (2). Despite the overall burden of HIV infection, women of reproductive age experience higher burden as compared to men. In Africa, the prevalence of HIV among women of reproductive age ranges from 1.4% to 6.7% (5).

Tanzania is ranked among the 22 priority countries constituting 90% of all pregnant women living with HIV worldwide (6). The country has one of the highest annual incidence of HIV infection in the world recording an incidence of 1.55 per 1000 in the year 2021 (7). About 36,700 women aged 15-49 were newly infected with HIV in 2020 and estimated 28,000 in 2021 (7,8). Although the country has significantly reduced the rate of mother-to-child transmission (MTCT) of HIV from 23% in 2011 to 11% in 2020, it is still behind the global targets and is one of the six countries with a slow pace or no decline in MTCT of HIV (6,9).

As one of the country with the highest HIV-incidence, Tanzania is a particular focus of the global fast-track approach to end HIV/AIDS as a public health threat by 2030. Driving the fast-track approach are the fast-track targets and speeding up implementation of effective HIV prevention and treatment services, and strategically allocating services and resources on areas and populations most affected. This can only be achieved through better understanding of the local epidemic.

The fast-track targets set for 2020 were 90-90-90 and 95-95-95 by 2030. That is by 2030, 95% of PLHIV should know their HIV-positive status, and 95% of people who know their status should be on treatment, and 95% of those on ART should attain and maintain viral suppression. Among adults living with HIV in 2016 in Tanzania, almost half (51.8%) were aware that they were HIV-infected, and only 47.1% were aware and on ART (10). Although women living with HIV were more likely to know their HIV-positive status and be on ART compared with the general population, the level of HIV status awareness and ART coverage was still very low. Only 55.4% of them were aware of their HIV-positive status and slightly over half (51.5%) of those infected were on ART (10). This hampers efforts to eliminate HIV/AIDS by 2030 (11,12).

For over two decades, the HIV sentinel surveillance (HSS) among pregnant women attending ANC have provided estimates of the burden and trend of HIV infections in Tanzania (13–16). Six rounds of HSS have been done since 2000 and results indicate a decline in the prevalence of HIV infections among pregnant women in Tanzania from 9.6% in 2001 to 5.6% in 2011 (15). However, results from the last HSS, in 2017 (unpublished), showed an increase in prevalence to 6.1%.

The HSS also plays a vital role in informing the needs for HIV testing, and other HIV and STIs-related needs at the national and sub-national levels to prevent mother-to-child transmission (PMTCT) of HIV (13,17). The HSS has also been expanding to include all regions, representing facilities from urban, semi-urban, and rural areas. In line with the ambitious global targets to, recent surveys also collected information on HIV status awareness and ART coverage among pregnant women which help to inform on the progress towards the goals and effectiveness of the interventions.

In line with the need to close the gap in HIV control, essential to the national and global response to HIV, we analysed data from the seventh HSS round conducted in 2020 to update national data on the burden of HIV, awareness of HIV status, coverage of ART, and factors associated with HIV infection among pregnant women in Tanzania mainland.

## Materials and methods

### Surveillance duration and site selection

Data for the seventh HSS was collected from September to December 2020 in 26 regions of Tanzania’s mainland from 159 antenatal clinics (ANCs) providing PMTCT services. ANC coverage of at least one visit is more than 98% and PMTCT services are integrated into ANCs (9). In 2019, 2.2 million pregnant women accepted HIV testing, and 98.3% of all pregnant women attended PMTCT services (9). Pregnant women with HIV-positive test results are initiated on ART irrespective of their CD4 cell count, in line with the national guidelines (18). Women who opt out at their first ANC visit are informed that they can access HIV testing at any future visit if they change their minds (18). As for the previous HSS rounds, two urban, two semi-urban, and two rural sites were selected from each region, except for the Dar es Salaam region, where all six sites are classified as urban.

### Survey population

All pregnant women aged 15 years and above attending ANC on their first visit in the current pregnancy during the survey period were eligible for inclusion in the ANC HSS. However, only those who provided written consent and were willing to give a blood sample for routine PMTCT HIV testing were included in the surveillance. To achieve the targeted sample size, all sites carried out data collection continuously for three consecutive months.

### Data collection

This surveillance was integrated into routine ANC activities where all women attending the ANC on a particular day were informed of the ongoing surveillance. The provider explained the surveillance procedures and obtained written informed consent from the participant for each component of the survey (the interview and routine rapid testing for HIV). In addition, a non-identifying unique survey ID code (HSS number) pre-printed on barcode stickers was assigned to each prospective survey participant found to be eligible.

The surveillance questionnaire was developed in English, translated into Swahili, and then back-translated into English by a third party to verify the accuracy of the translation. A trained RCH provider, using an electronic tablet loaded with open data kit (ODK), a data collection software, administered the questionnaire. It captured the HSS number, ANC number, age, marital status, gravidity, education level, residence, employment, HIV status, ART use, and HIV test results. Electronic data were uploaded to a password-protected server at the end of each day.

### HIV testing procedures

Following the interview, all participants, including those who knew their HIV status, were offered HIV counseling and testing according to the national guidelines and the national rapid testing algorithm (19). The provider conducted HIV and syphilis testing using rapid SD Bioline HIV/ Syphilis Duo test kits, followed by a Unigold test to confirm any HIV-positive results. The SD Bioline HIV/ Syphilis Duo test kit offers similar sensitivity and specificity for HIV as the single HIV SD Bioline (20–23). This allows for comparability with the results of the previous rounds of this surveillance. The Ministry of Health (MOH) adopted the kit owing to its cost-effectiveness in testing for both HIV and syphilis and enables broader syphilis testing coverage (21,22). Each participant received their result in a post-test counseling session and was referred to HIV prevention, care, treatment, and support services as appropriate. Participants with positive syphilis test results were treated according to National STI Management Guidelines (24). HIV test result data were recorded on a register and entered into an ODK form.

### Training of research personnel

A group of national-level facilitators was trained on implementing the ANC HSS survey by the principal investigator and the co-investigators. These national-level facilitators then conducted zonal training workshops organized in five zones before the commencement of the survey. Participants included nurses attending ANC clients from all 159 sites and regional and district personnel involved in the survey implementation. The training included a review of operational procedures and field protocols for ANC HSS based on routine data. It also included a training on the updated ANC HSS data collection form and HIV and syphilis testing procedures. In addition, a team of national-level study staff was trained to monitor the quality of routinely collected PMTCT data and the quality of PMTCT HIV and syphilis testing.

### Data management and analysis

Sociodemographic data and routine HIV testing data collected were uploaded daily to a password-protected cloud storage system by each survey team member to allow for real-time data monitoring. To minimize data entry errors, questions in the ODK had prompts and checks for data validation and correction of identified errors.

Analysis was conducted using Stata Statistical software ver. 17 (25) (College Station, TX: Stata Corp LLC.). The proportion of women who were infected with HIV was calculated at the national and regional levels and was also disaggregated by demographic characteristics. These estimates, and the logistic regression estimates, considered the survey design of this surveillance (clustering at the health facility level and stratification by locality - rural, semi-urban, and urban). The estimates were also weighted using inverse probability weights based on the total number of women who attended ANC during the study period at each region. To compare with previous surveys, crude estimates (unweighted and not considering the survey design) of HIV infection at the national level were calculated and presented. Multivariable logistic regressions were used to examine the independent association between HIV infection and various independent variables including age, marital status, parity, education level, source of income, and locality. All the variables that attained a p-value <0.2 in the univariable analysis were included in the multivariable models. The level of statistical significance was set at p<0.05.

We also estimated the proportion of HIV-positive status awareness among women living with HIV in the survey and the ART coverage among those aware of their HIV-positive status. These estimates were disaggregated by regions and by demographic characteristics of the women. Because the HIV status awareness and ART coverage were self-reported, we conducted and presented the sensitivity analysis of their estimates. For ART coverage, we estimated and presented estimates where 10% of those who reported being aware of their status but not on ART were considered to be on ART, and 5% of those who reported being aware of their HIV status and on ART were considered to be not on ART (10). And for HIV-positive status awareness, 10% of those who reported being unaware of their HIV-positive status were considered to have previously been diagnosed. These estimates were derived from the 2016/17 Tanzania HIV Impact survey (10).

### Ethical consideration

Study staff sought informed consent from potential participants before they engaged them in the survey. Pregnant women who did not consent to participate in the survey received routine ANC care as supposed. Participants were assured that their responses would be kept confidential and that no harm would come to them because of agreeing or not agreeing to participate in the survey. Approval to conduct this study was provided by the MUHAS Institutional Review Board (MUHAS-REC-07-2020-298). Strict protection ensured data security and confidentiality. The ANC HSS survey did not collect any personally identifiable information other than what was essential to meet the assessment objectives.

## Results

### Demographic characteristics

Of the 39,516 pregnant women included in the 2020 ANC HSS surveillance, 38,783 (98.1%) consented to participate in the surveillance and had their HIV results. The median age (IQR) of the participants was 25 (21–30) years, and a third of them (33.2%) were aged between 20-24 years. Over three-quarters of the participants (78.5%) reported being married, and 61.0% had attained primary school education. About half of the participants (48.8%) were housewives, while 44.5% had at most two previous pregnancies, and a quarter of all participants (26.3%) were primigravids. Almost half of the participants, 48.2%, were recruited from facilities located in urban areas —**Table 1**.

**Table 1:** Prevalence of HIV by demographic.

| Variable | Total | HIV Infection |
| --- | --- | --- |
| Age group (years) | N (%) | Prevalence (%) (95% CI) |
| 15-24 | 17,766 (45.8) | 3.0 (2.6- 3.4) |
| 25-34 | 15917 (41.0) | 7.4 (6.6- 8.2) |
| 35-44 | 4746 (12.2) | 11.2 (9.9- 12.7) |
| 45+ | 354 (0.9) | 18.2 (12.1- 26.4) |
| **Education level** |  |  |
| None | 4,896 (12.6) | 7.2 (6.2-8.4) |
| Primary | 23,627 (60.9) | 6.8 (6.1-7.5) |
| Secondary | 9,035 (23.3) | 4.9 (4.1-5.9) |
| Post-secondary | 1,225 (3.2) | 4.7 (3.5-6.1) |
| **Marital status** |  |  |
| Single | 2,835 (7.3) | 10.4 (8.7-12.4) |
| Divorced/separated | 308 (0.8) | 22.4 (16.3-29.9) |
| Widowed | 113 (0.3) | 23.0 (16.5-31.2) |
| Co-habiting | 5,081 (13.1) | 6.8 (5.9-8) |
| Married | 30,446 (78.5) | 5.6 (5.1-6.3) |
| **Employment status** |  |  |
| Employed | 2,014 (5.2) | 7.7 (6.2-9.5) |
| Housewife | 18,956 (48.9) | 5.4 (4.9-6) |
| Self-employed | 15,032 (38.8) | 7.5 (6.6-8.5) |
| Unemployed | 2,781 (7.2) | 5 (3.7-6.7) |
| **Total number of previous pregnancies** |  |  |
| 0 | 10,199 (26.3) | 2.7 (2.3-3.1) |
| 1-2 | 17,285 (44.6) | 7.1 (6.4-7.8) |
| 3-4 | 7,916 (20.4) | 9.6 (8.6-10.8) |
| >4 | 3,383 (8.7) | 6.0 (5.1-7.0) |
| **Locality** |  |  |
| Rural | 6,925 (17.9) | 4.3 (3.4-5.3) |
| Semi-urban | 13,020 (33.6) | 7.3 (6-8.9) |
| Urban | 18,838 (48.6) | 6.4 (5.6-7.3) |
| **Health facility level** |  |  |
| Dispensary | 6,131 (15.8) | 5.5 (4.3-6.9) |
| Health Centre | 23,837 (61.5) | 5.9 (5.2-6.8) |
| Hospital | 8,815 (22.7) | 8.0 (6.7-9.7) |

### Prevalence of HIV by demographic characteristics of participants

The overall prevalence of HIV infection among first-visit ANC attendees in Tanzania mainland was 5.9% (95%CI: 5.3% - 6.6%). The prevalence of HIV was highest among women aged 45 years and above (18.2%), among widowed women (23.0%), and divorced/separated women (22.4%). Women in semi-urban areas were observed to have a higher prevalence (7.3%) compared to those in urban (6.4%) and rural areas (4.3%). Women in their first pregnancy had the lowest prevalence compared to those who had pregnancy before —**Table 1**. The crude prevalence of HIV was 6.3% (95%CI: 6.0%- 6.5%).

### Prevalence of HIV infection by regions

The prevalence of HIV infection was unvenly distributed ranging from the lowest in the Manyara region (2.0%) to the highest in Njombe region (16.4%) —**Table 2**.

**Table 2:** Prevalence of HIV infection by regions.

| Region | Total<br>N | HIV Infection<br>n | %, (95% CI) |
| --- | --- | --- | --- |
| Arusha | 1,542 | 51 | 3.3 (1.7-6.4) |
| Dar es Salaam | 2,874 | 167 | 5.8 (4.3-7.9) |
| Dodoma | 1,749 | 68 | 3.9 (3.5-4.3) |
| Geita | 3,460 | 231 | 6.7 (5.6-8) |
| Iringa | 882 | 120 | 13.6 (10.9-16.9) |
| Kagera | 1,481 | 73 | 4.9 (3.2-7.5) |
| Katavi | 1,224 | 72 | 5.9 (4.3-8.1) |
| Kigoma | 1,372 | 29 | 2.1 (1.1-4.1) |
| Kilimanjaro | 968 | 68 | 7 (4.3-11.2) |
| Lindi | 508 | 36 | 7.1 (5.7-8.8) |
| Manyara | 924 | 18 | 1.9 (1.0-3.6) |
| Mara | 1,509 | 83 | 5.5 (2.8-10.5) |
| Mbeya | 2,032 | 241 | 11.9 (9.7-14.4) |
| Morogoro | 728 | 35 | 4.8 (2.2-10.4) |
| Mtwara | 814 | 39 | 4.8 (3.9-5.9) |
| Mwanza | 2,232 | 128 | 5.7 (4-8.1) |
| Njombe | 670 | 110 | 16.4 (11.2-23.5) |
| Pwani | 1,841 | 149 | 8.1 (3.8-16.5) |
| Rukwa | 1,265 | 80 | 6.3 (4.6-8.7) |
| Ruvuma | 1,119 | 105 | 9.4 (4.6-18.2) |
| Shinyanga | 1,477 | 102 | 6.9 (3.9-11.8) |
| Simiyu | 1,956 | 85 | 4.3 (2.7-6.8) |
| Singida | 1,549 | 67 | 4.3 (2.4-7.8) |
| Songwe | 1,902 | 132 | 6.9 (5.3-9.1) |
| Tabora | 1,408 | 96 | 6.8 (4.6-10.1) |
| Tanga | 1,297 | 71 | 5.5 (4.4-6.8) |
| **Total** | **38,783** | **2456** | **5.9 (5.3 – 6.6)** |

### Factors associated with HIV infection among pregnant women

**Table 3** shows the results of logistic regression analyses. Odds of being HIV positive increased with age (trend z= 26.02, p<0.001). Pregnant women aged 25-34 years had 2.2 times higher odds of having HIV than pregnant women aged 15-24 years (aOR=2.2, 95% CI: 1.9-2.5). Similarly, women aged 35-44 years (aOR=3.9, 95% CI: 3.2-4.6), and women of 45 years and above (aOR=7.0, 95% CI: 4.4-11.1) had respectively almost 4 and 7 times higher odds of having HIV infection compared to pregnant women aged 15-19 years (**Table 3**). The odds decreased with an increase in education level (trend z=-4.54, p<0.001).

**Table 3:** Factors associated with HIV infection among pregnant women.

| Variable | Positive n (%) | cOR (95% CI) | aOR (95% CI) | p-value |
| --- | --- | --- | --- | --- |
| **Age group (years)** |  |  |  |  |
| 15-24 | 582 (3.3) | Ref | Ref |  |
| 25-34 | 1246 (7.8) | 2.6 (2.3-2.8) | 2.2 (1.9-2.5) | <0.001 |
| 35-44 | 557 (11.7) | 4.1 (3.5-4.7) | 3.9 (3.2-4.6) | <0.001 |
| 45+ | 71 (20.1) | 7.2 (4.4-11.7) | 7.0 (4.4-11.1) | <0.001 |
| **Education level** |  |  |  |  |
| None | 353 (7.2) | 1.6 (1.2-2.2) | 2.3 (1.6-3.4) | <0.001 |
| Primary | 1599 (6.8) | 1.5 (1.1-2) | 1.9 (1.3-2.6) | <0.001 |
| Secondary | 447 (5.0) | 1.1 (0.8-1.5) | 1.2 (0.9-1.7) | 0.227 |
| Post-secondary | 57 (4.7) | Ref | Ref |  |
| **Marital status** |  |  |  |  |
| Single | 294 (10.4) | 1.9 (1.6-2.3) | 3.3 (2.7-4.0) | <0.001 |
| Co-habiting | 348 (6.9) | 1.2 (1-1.5) | 1.4 (1.2-1.7) | 0.001 |
| Divorced/separated | 69 (22.4) | 4.8 (3.2-7.2) | 4.8 (3.2-7.1) | <0.001 |
| Widowed | 26 (23.0) | 5 (3.2-7.7) | 3.4 (2.2-5.4) | <0.001 |
| Married | 1719 (5.7) | Ref | Ref |  |
| **Employment status** |  |  |  |  |
| Employed | 155 (7.7) | 1.6 (1.2-2.2) | 1.7 (1.1-2.4) | 0.008 |
| Housewife | 1032 (5.4) | 1.1 (0.8-1.5) | 1.2 (0.9-1.7) | 0.262 |
| Self-employed | 1131 (7.5) | 1.6 (1.1-2.1) | 1.6 (1.1-2.2) | 0.012 |
| Unemployed | 138 (5.0) | Ref | Ref |  |
| **Total number of previous pregnancies** |  |  |  |  |
| 0 | 271 (2.7) | Ref | Ref |  |
| 1-2 | 1219 (7.1) | 2.8 (2.4-3.2) | 2.2 (1.9-2.6) | <0.001 |
| 3-4 | 763 (9.6) | 3.9 (3.3-4.6) | 2.2 (1.8-2.6) | <0.001 |
| >4 | 203 (6.0) | 2.3 (1.9-2.9) | 1.1 (0.8-1.4) | 0.479 |
| **Locality** |  |  |  |  |
| Rural | 296 (4.3) | Ref | Ref |  |
| Semi-urban | 954 (7.3) | 1.8 (1.3-2.4) | 1.5 (1.0-2.2) | 0.030 |
| Urban | 1206 (6.4) | 1.5 (1.1-2.1) | 1.5 (1.1-2.1) | 0.012 |
| **Health facility level** |  |  |  |  |
| Dispensary | 335 (5.5) | Ref | Ref |  |
| Health Centre | 1412 (5.9) | 1.1 (0.8-1.5) | 1.1 (0.8-1.4) | 0.669 |
| Hospital | 709 (8.0) | 1.5 (1.1-2.1) | 1.4 (0.9-2.0) | 0.099 |

Marital status was also a significant predictor of HIV infection among women in this survey. Women who were widowed were 3.4 times more likely to test HIV positive compared to those who were married (aOR=3.4, 95%CI: 2.2-5.4). Similar findings were observed for those who were divorced/separated (aOR=4.8, 95%CI): 3.2-7.1). Women who were single were three times more likely to test positive for HIV compared to those who were married (aOR=3.3, 95%CI: 2.7-4.0). Compared to women in rural areas, women residing in urban and suburban areas had 50% higher odds of HIV infection. **Table 3**.

In the sensitivity analysis of the HIV status awareness, the HIV status awareness was 73.7% when 10% of HIV-positive women who reported being unaware of their HIV-positive status were considered to be aware of their HIV-positive status. And ART coverage was 87.1% in the sensitivity analysis when 10% of those who reported being aware of their HIV-positive status but not on ART were considered to be on ART, and 5% of those who reported being aware of their HIV status and on ART were considered to be not on ART.

### Awareness of HIV positive status and ART coverage

The estimated HIV status awareness among women who tested positive was 70.9% (95% CI: 67.5%-74.0%). The estimated ART coverage among those aware of their status was 91.6% (86.5%-94.9%). Overall, 66.6% (95% CI: 62.4%- 70.6%) of all women who tested positive for HIV in this survey knew their HIV status and were on ART—**Fig 1**.

**Fig 1:**
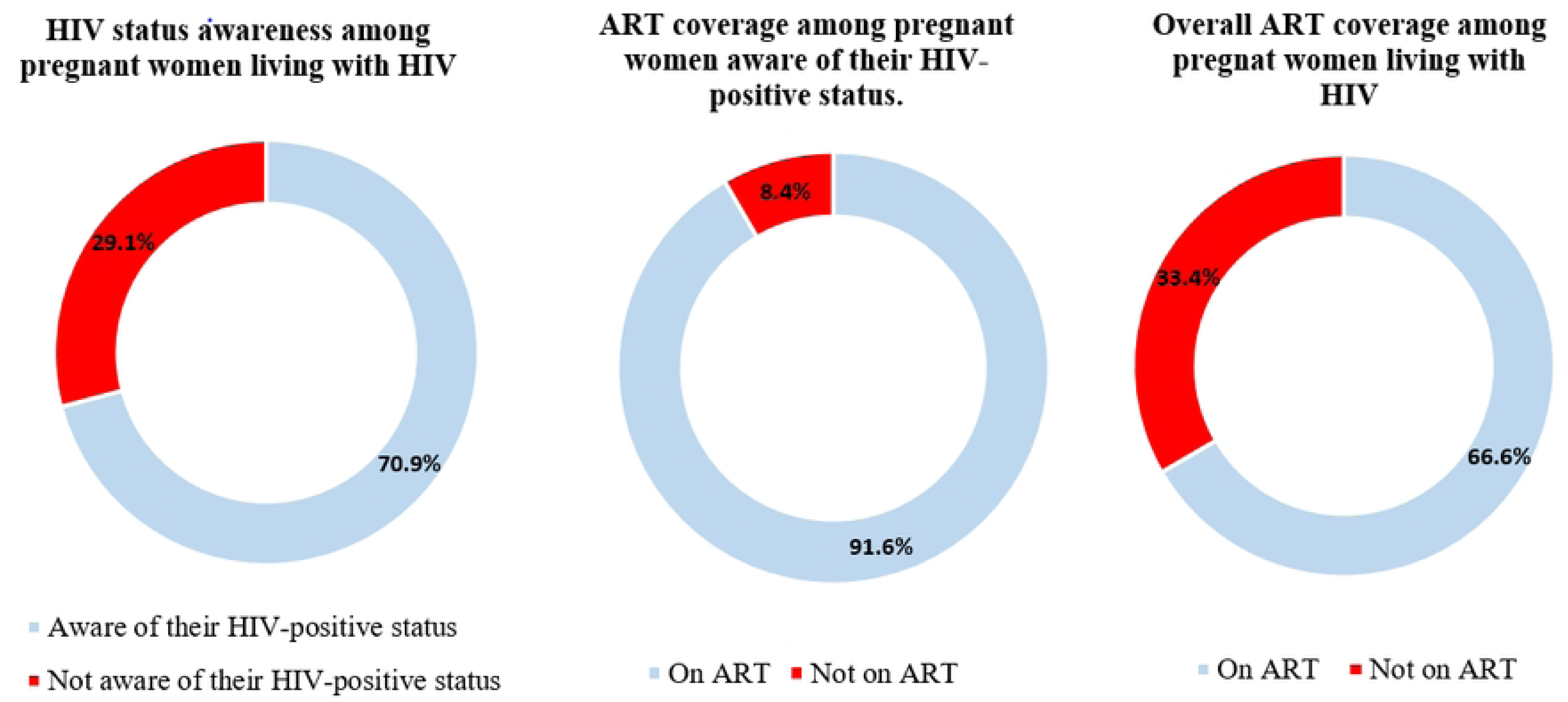
Progress on the global fast-track targets among pregnant women attending ANC in Tanzania mainland.

### Awareness of HIV Positive Status and ART coverage by regions

None of the regions has attained the first 95% target. The proportion of HIV-positive pregnant women attending ANC who knew their status varied among regions ranging from 92% in Ruvuma to 44% in Manyara and 47% in Katavi —**Fig 2**.

**Fig 2:**
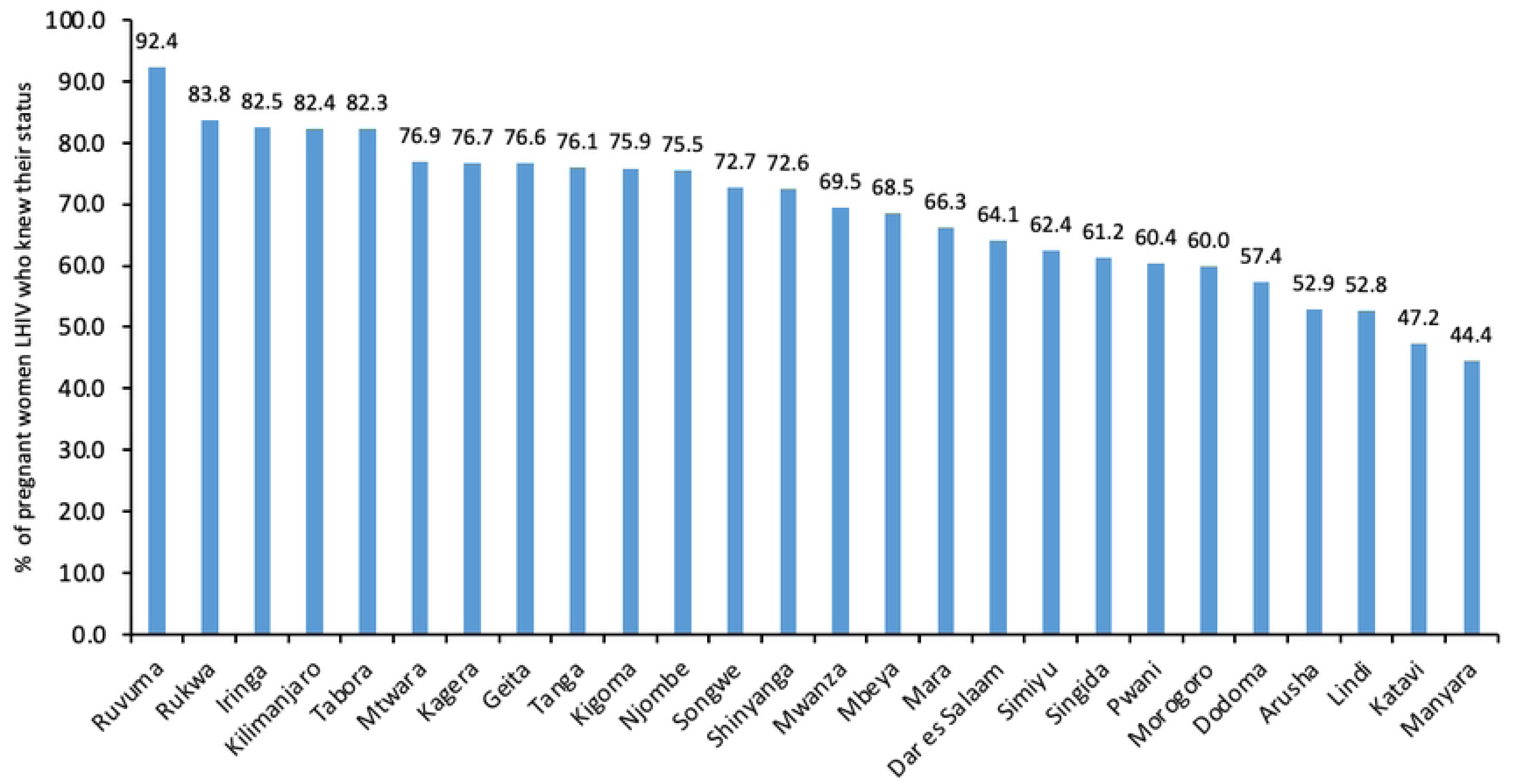
Percentage of HIV-positive women attending ANC who knew their status by region.

Eleven regions had already attained the second 95% target. These were Kigoma, Morogoro, Shinyanga, Simiyu, Tanga, Kagera, Mara, Mbeya, Arusha, Singida, and Katavi. Three regions, Pwani (51%), Manyara (75%), and Mtwara (77%), had the lowest ART coverage. Dar Es Salaam (87%), Geita (86%), Lindi (89%), Njombe (83%), and Rukwa (87%) had ART coverage below 90% among women living with HIV who knew their status —**Fig 3**.

**Fig 3:**
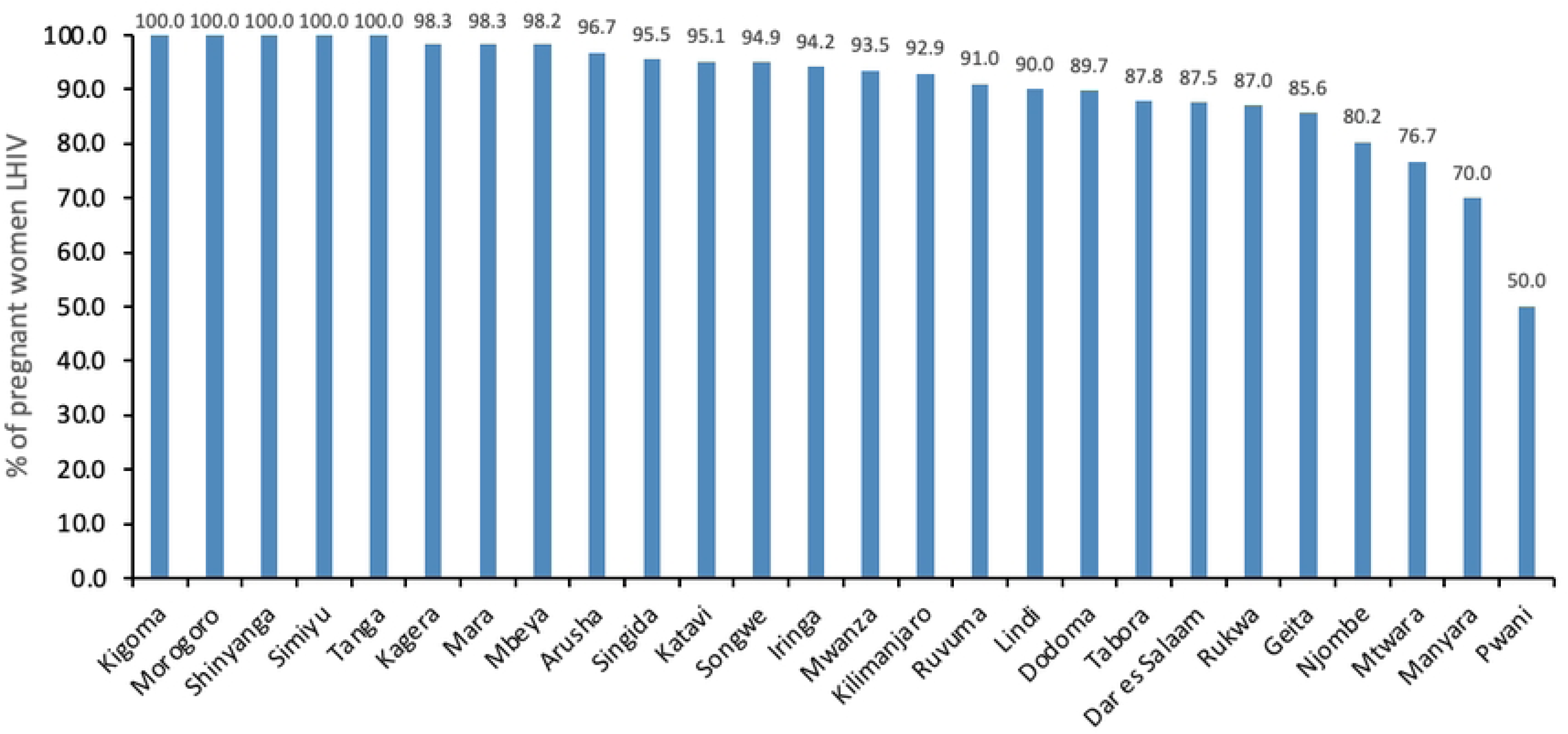
Percentage of HIV-positive women attending ANC who knew their status that were on ART by region.

### HIV Status awareness and ART coverage by demographic characteristics

Among pregnant women who tested positive for HIV, the HIV-positive status awareness was lowest (56.6%) among women aged 15-24 years, while the highest (80.4%) was among women aged 35-44 years. Less than half (46.6%) of primigravids who tested positive were aware of their HIV-positive status while over 70% of women with at least one previous pregnancy were aware of their HIV-positive status. Three-quarters (75.5%) of the pregnant women in semi-urban areas were aware of their HIV-positive status whereas, in comparison, two third of those residing in the rural (66.4%) and urban areas (68.2%) were aware of their HIV-positive status. ART coverage was highest among rural (97.1) resindents and lowest among semi-urban dwellers (88.1%). ART coverage was lowest (30%) among pregnant women of 45 years and above. Women with secondary (85.5%) or post-secondary education (86.0%) had ART coverage lower than 90% . Primigravidshad the lowest ART coverage (80.0%) compared to women with at least one previous pregnancy who had over 90% coverage—**Table 4**.

**Table 4:** HIV Status awareness and ART coverage by demographics.

| Variable | Total Positive | Aware Status % (95% CI)<br>N=2456 | On ART % (95% CI)<br>N=1708 |
| --- | --- | --- | --- |
| **Age group (years)** |  |  |  |
| 15-24 | 582 | 56.6 (51.1- 62.0) | 90.9 (85.9- 94.3) |
| 25-34 | 1246 | 73.1 (69.1- 76.7) | 92.9 (89.3- 95.4) |
| 35-44 | 557 | 80.4 (75.7- 84.3) | 94.3 (88.9- 97.1) |
| 45+ | 71 | 68.8 (59.5- 76.8) | 30.3 (05.3- 77.0) |
| **Education level** |  |  |  |
| None | 353 | 69.7 (63.4- 75.4) | 92.8 (86.2- 96.4) |
| Primary | 1599 | 72.1 (68.6- 75.4) | 93.1 (89.7- 95.5) |
| Secondary | 447 | 69.1 (63.2- 74.5) | 85.5 (71.2- 93.3) |
| Post-secondary | 57 | 55.6 (37.5- 72.3) | 86.0 (68.1- 94.6) |
| **Marital status** |  |  |  |
| Single | 294 | 72.7 (59.0- 83.1) | 91.0 (62.5- 98.4) |
| Co-habiting | 348 | 64.9 (58.3- 71.1) | 94.9 (89.7- 97.5) |
| Married | 1719 | 72.1 (68.4- 75.5) | 91.0 (84.1- 95.0) |
| Divorced/separated | 69 | 72.3 (64.9- 78.6) | 91.7 (84.2- 95.8) |
| Widowed | 26 | 57.3 (33.5- 78.1) | 90.9 (71.0-97.6) |
| **Total number of previous pregnancies** |  |  |  |
| 0 | 271 | 46.6 (39.7- 53.6) | 80.0 (66.6- 88.9) |
| 1-2 | 1219 | 70.0 (66.0- 73.6) | 90.2 (84.1- 94.1) |
| 3-4 | 763 | 78.5 (74.6- 81.9) | 95.6 (91.7- 97.8) |
| >4 | 203 | 81.6 (74.7- 86.9) | 93.9 (87.3- 97.1) |
| **Locality** |  |  |  |
| Rural | 296 | 66.4 (59.9- 72.3) | 97.1 (91.9- 99.0) |
| Semi-urban | 954 | 75.5 (70.0- 80.4) | 88.1 (76.2- 94.5) |
| Urban | 1206 | 68.2 (63.3- 72.8) | 93.4 (89.3- 96.0) |

## Discussion

HIV infection was prevalent among 5.9% of pregnant women attending ANC in Tanzania and with a marked regional variation. This study further revealed that Tanzania is still far behind in attaining the first fast-track targets among pregnant women. HIV status awareness among pregnant women in this survey was only at 70.9%, also with a wider regional variability. Among pregnant women who knew their HIV status, 91.6% were on ART. Age, marital status, education level, number of pregnancies, and locality were significant predictors of HIV infection among pregnant.

Evidence elsewhere have been consistent on a higher burden of HIV infection among pregnant women compared to the general population (17,26). In this survey, the observed prevalence of HIV infection among pregnant women in Tanzania mainland was also higher (5.9%) compared to that of the general population (4.7%). However, the prevalence is lower compared to that of women reported in the population-based survey in 2017 of 6.4% (10). Prevalence of HIV infection among pregnant women aged 15-49 in Tanzania mainland from 2003/04 and 2006/07 HSS were higher than those of women from the parallel population-based surveys (14,27–30). During the 2011 and 2017 HSS however, the estimates were lower than those reported in the parallel population surveys (15,27,30,31). The trendy decline in prevalence in the recent surveys over the years may reflect ART-associated survival of women living with HIV. This is more pronounced in the population-based surveys than in the surveillance of pregnant women because older women living with HIV have lower fertility than women not living with HIV (32–34)(34). It is also worthnotting that although fertility among women living with HIV has improved in Tanzania, it is still lower than that among women not living with HIV, especially among older women (35). Therefore, the prevalence of HIV among pregnant women is slightly lower than that of women in the general population. Estimating HIV prevalence for women in the general population using the HSS findings should therefore be interpreted with caution.

To campare the prevalence of HIV infection among pregnant women across rounds of HSS, we used the crude estimate. The crude prevalence in this survey was 6.3%, which is higher compared with the prevalence of the preceeding survey of 6.1% reported in 2017. From the first round of HSS in 2000 until the fifth round of HSS in 2011, the HIV prevalence had been declining from 10% to 5.3% (15). The incline in prevalence in the 2017 and 2020 surveys coincides with the introduction of Option B+ in Tanzania in September 2013, that recommended to initiate ART to pregnant women regardless of the CD4 count(14,15,36). Option B+ is an intervention behind the global reduction of MTCT (10,12,37,38). In this survey, a large proportion of HIV infections among pregnant women,70%, is actually among women who knew their status and were not newly diagnosed.

Similar to the previous ANC surveys and other population-based surveys, the 2020 ANC HSS observed wide regional variations in HIV prevalence ranging from 1.9% in the Manyara region to 16.4% in the Njombe region. Iringa region (13.6%) and Mbeya region (11.9%) ranked second and third for HIV prevalence, in line with findings from the previous surveys(10) (30). The persistently high burden in these regions could be partly explained by the continued efforts by the Government of Tanzania and its stakeholders to address AIDS-related deaths resulting in improved survival for people living with HIV infection. However, these regions still had low HIV status awareness which may indicate the potential of high transmission rate. High prevalence, low HIV status awareness, and subsequently low ART coverage may fuel onward transmission and increase incidence of HIV infections (39).

The overall HIV status awareness has increased from 43% in 2017 to 70% in 2020, but it is still below 95% target. Only one region (Ruvuma) reached 90% in this survey. Although 91.6% of pregnant women who were aware of their HIV-positive status were on ART, overall only 66.6% of all women living with HIV in this survey were diagnosed and on ART. Closing this gap in HIV testing and treatment is a critical part of the response to the HIV epidemic as it has significant implications for preventing onward transmission of HIV and its impact on the incidence at the population level (40). Tanzania, which is also among the countries with the highest HIV incidences globally, urgently needs to strengthen the existing HIV testing services (HTS) and employ new and innovative ways to get people tested for HIV. As revealed in this study, the expansion of HTS also needs to be done strategically by prioritizing regions and population groups with lower coverage of HTS and ART, such as young women and adolescents. Coverage was almost twice for non-primigravida compared to primigravida women indicating that integration of HTS into other services increases the uptake of HTS which is inline with the findings and recommendation fom other studies (41–43). These should go hand in hand with linkage to treatment and care (3). Efforts should be directed towards the most vulnerable areas with high prevalence, low HIV status awareness, and low ART coverage (44). It is relatively more challenging, logistically and financially, to determine incidences, but data from surveillances such as this one may help inform on the incidence and implement interventions. Additionally, through surveys such as this, a score for predicting incidence using the data from surveys may be developed and validated.

The analysis of factors associated with HIV determined that older age and lower level of education were associated with higher HIV prevalence. Age factor reflects the increased survival of PLHIV in the era of ART. Since this was a cross-sectional survey, it is not surprising that older pregnant women were more likely to be found with HIV infection than their younger counterparts. A cross-sectional survey, by nature of its design, studies survivors of a chronic infection like HIV and hence high prevalence amongst them. The prevalence of HIV decreased with an increase in education level like in the population-based HIV survey in Tanzania (10). Education attainment delays sexual debut, keep girls in school which offers protective environment for unwanted pregnancies and risky sexual behaviors (45). Evidence in the current survey is contrary to the previous surveys, where the prevalence of HIV was higher among people with higher levels of education (14). This was thought to be attributed by lack of access to HIV preventive methods and their positions in society rendered them at risky sexual behaviours (14). In the current survey, pregnant women in marital union had the lowest prevalence of HIV infection (5.7%) compared to those outside of marital union. The highest prevalence was observed among pregnant women reported to be widowed (23.0%), divorced or separated (22.4%), and those who were single (10.4%). The relationship between marital status and HIV infection is complex and largely understudied but cultures and sexual behaviours have a large role in this interplay (46,47). Consistent with our findings, the recent population demographic survey indicated a lower lifetime number of sexual patners among married women compared to women outside marital union. Married women are also more likely to use condom during sexual intercourse with a person who is not their husband or living with compared to unmarried women (48), preventing them from new HIV infections.

Living in urban and semi-urban areas was associated with higher HIV infections compared to living in rural areas in this survey. Similar pattern has been observed among pregnant women over the years in Tanzania (30). The sexual behavioral differences of women in the urban and rural may influencing this observation. Women in rural areas have a lower average number of lifetime sexual partners and fewer have sexual intercourse outside of marital union or with a non-cohabiting partner compared to women in the urban areas (48). However the proportion of condom use in these instances are comparable in the two settings (48).

It has been observed in several studies that individuals who are affected with HIV have lower participation in the labor force particularly in the manual labors and understandably, severity of disease is associated with lower employment (49,50). Nevertheless in this national wide survey, the prevalence of HIV was higher among those who reported to be employed or self-employed. This observation may be a result of survival and stability of those who are employed compared to those who are not employed. People affected with HIV who are employed are likely to test for HIV, diagnosed earlier, and initiate ARTs with better adherence than people who are not employed(49,51,52). This can in turn translates to better survival and stable health among those employed compared to those who are not employed (51,52). Employment increases financial and economic empowerment among women, which may increase access to health care services, reduce HIV-related morbidity and mortality, and improve the quality of life for PLHIV (51,53).

## Conclusion

One in every twenty pregnant women attending ANC in Tanzania is infected with HIV. Prevalence varied between regions, but HIV is especially prevalent among pregnant women who are older, those with lower or no formal education, those outside marital union and those living in urban and semi-urban areas. Only seven out of ten pregnant women living with HIV are aware of their HIV-positive status. However, ART is covered in about nine out of ten pregnant women living with HIV who were aware of their HIV-positive status. Behind the global fast-target to end HIV/ AIDS, about a third of pregnant women living with HIV initiating ANC were not on ART. Interventions to increase HIV testing and linkage to care among women of reproductive age should be intensified.

## Data Availability

We cannot at this time share the de-identified data file with the third party owing to the Tanzania Health Research Regulation that requires the third party to sign the Data Transfer Agreement. Data may be shared upon request by any researcher to the Ministry of Health (the owner) and upon fulfilling the regulatory requirements.

## Acknowledgment

We wish to thank all regional and district medical officers, regional and district reproductive, and sexual health coordinators for their support in the implementation of the survey. Our sincere appreciations to the nurses at the facilities for their commitment throughout the study. BSJ passed away before the submission of the final version of this manuscript. EM (Corresponding author) accepts responsibility for the integrity and validity of the data collected and analyzed.

